# Investigating the effects of absolute humidity and human encounters on transmission of COVID-19 in the United States

**DOI:** 10.1101/2020.10.30.20223446

**Authors:** Gary Lin, Alisa Hamilton, Oliver Gatalo, Fardad Haghpanah, Takeru Igusa, Eili Klein, For the CDC MInD-Healthcare Network

## Abstract

**Background:** Mounting evidence suggests that the primary mode of transmission of SARS-CoV-2 is aerosolized transmission from close contact with infected individuals. Even though transmission is a direct result of human encounters, environmental conditions, such as lower humidity, may enhance aerosolized transmission risks similar to other respiratory viruses such as influenza.

**Methods:** We utilized dynamic time warping to cluster all 3,137 counties in the United States based on temporal data on absolute humidity from March 10 to September 29, 2020. We then used a multivariate generalized additive model (GAM) combining data on human mobility derived from mobile phone data with humidity data to identify the potential effect of absolute humidity and mobility on new daily cases of COVID-19 while considering the temporal differences between seasons.

**Results:** The clustering analysis found ten groups of counties with similar humidity levels. We found a significant negative effect between increasing humidity and new cases of COVID-19 in most regions, particularly in the period from March to July. The effect was greater in regions with generally lower humidity in the Western, Midwest, and Northeast regions of the US. In the two regions with the largest effect, a 1 g/m^3^ increase of absolute humidity resulted in a 0.21 and 0.15 decrease in cases. The effect of mobility on cases was positive and significant across all regions in the July-Sept time period, though the relationship in some regions was more mixed in the March to June period.

**Conclusions:** We found that increasing humidity played an important role in falling cases in the spring, while increasing mobility in the summer contributed more significantly to increases in the summer. Our findings suggest that, similar to other respiratory viruses, the decreasing humidity in the winter is likely to lead to an increase in COVID-19 cases. Furthermore, the fact that mobility data were positively correlated suggests that efforts to counteract the rise in cases due to falling humidity can be effective in limiting the burden of the pandemic.

## Introduction

To date, the coronavirus disease 2019 (COVID-19) pandemic has claimed over 225,000 lives in the United States alone, with more than 8.6 million confirmed cases (*1*). The primary mode of transmission of the severe acute respiratory syndrome coronavirus 2 (SARS-CoV-2) that causes COVID-19 is close contact with infected individuals (*2, 3*). Growing evidence suggests aerosols (*4, 5*), defined as particulates less than 5 µm in diameter (*6, 7*), likely play the most important role in transmission (*8*). The first surge of cases in the United States was seen in the late winter of 2020, but the vast majority of the epidemic to date has occurred during the spring and summer. As the fall season begins in most areas of the northern hemisphere, the weather will trend towards colder and drier. Falling levels of absolute humidity have been shown to increase transmission rates of other respiratory viruses, such as influenza (*9*), posing significant concern regarding potential increases in the number of COVID-19 cases in the fall and winter.

While several studies have suggested there may be a relationship between climatic factors such as temperature and/or humidity and COVID-19 (*10*–*16*), uncertainty still remains in the precise environmental and biological mechanisms (e.g., relative humidity, vapor pressure, and temperature) behind aerosol and droplet transmissions and viral survival of SARS-CoV-2 (*17*). In influenza, lower atmospheric moisture has been shown to increase the production of aerosol nuclei and viral survival time (*9*), which translates to higher risks of airborne transmission. Other climatic factors that may impact transmission include temperature and air quality (*18, 19*); however, absolute humidity can still provide a surrogate measure for indoor air moisture and temperature (*20*).

Initial efforts to slow the spread of COVID-19 focused on reducing contacts between individuals through social-distancing measures such as large-scale lockdowns, which were significantly associated with reductions in cases (*21*). However, as the initial lockdowns were lifted and movement of individuals increased, the correlation between mobility and case growth rates weakened (*22*). Though some counties and states saw increases in cases, others saw decreases without corresponding increases in movement by any metric. Thus, other factors besides mobility patterns are likely to be drivers of transmission.

Analyses of the factors influencing COVID-19 have used either climate data (*19, 23*–*25*) or human mobility data (*21*), but no study that we know of has considered changes in both climate and human mobility on COVID-19 outbreaks in the United States. Preliminary studies have investigated these effects in China but did not consider the varying sensitivities to humidity for different climatological regimes, which can lead to weak detection in humidity impacts on transmission (*26*). Understanding the potential for climatic factors to increase transmission in the fall and winter is crucial for developing policies to combat the spread of the SARS-CoV-2. While the interaction between environmental factors and human encounters is complex, accounting for this relationship is necessary for deciding business and school reopenings. Furthermore, as indoor gatherings, which typically increase in frequency and size in the winter, are one of the largest risk factors for transmission (*7, 27*), greater understanding is needed as to the added risk of changes in weather to aid decisions on when to restrict gatherings or implement mandates for protective face coverings. In this study, we assess the relative impact of absolute humidity in different climatological regimes and human mobility on reported cases of COVID-19 across counties in the US.

## Methods

### Data sources

Daily average absolute humidity for each US county, excluding territories, was calculated using temperature and dewpoint data from the National Centers for Environmental Information (*28*) at the National Oceanic and Atmospheric Administration (NOAA). Time series data for the year 2020 from US weather stations were acquired from the NOAA Global Summary of the Day Index (*29*). Weather stations were mapped using latitude and longitude to corresponding counties using the Federal Communications Commission (FCC) Census Block API (*30*). For counties without a weather station, we used data from the nearest station, which was calculated based on distance from the county’s spatial centroid using the haversine formula. For counties with multiple stations, data were averaged across all stations in a county. Absolute humidity was calculated using average daily temperature and average daily dew point (see (*31*)).

Data on visits to non-essential businesses from March 10, 2020 to September 29, 2020 was obtained from the Unacast Social Distancing Scorecard (*32*). We specifically utilized the metric that measures visits to non-essential places by comparing the rate on the county-level to the national average. This temporal measure allowed us to compare across counties. The data on non-essential visits was calculated as the percent difference from before policy interventions (e.g., shelter-in-place orders) began to impact movement.

Confirmed case data were extracted from the Johns Hopkins Center for Systems Science and Engineering (*1*), and the population size data for each county were obtained from the US Census Bureau (*33*) for 3,137 counties from March 10, 2020 to September 29, 2020. Daily cases were obtained from the confirmed case count by taking a simple difference between the days. Any data incongruencies, such as negative case counts, were omitted in our analysis.

### Statistical Analysis

The United States is geographically large, and the timing and magnitude of changes in absolute humidity can vary widely across regions. In order to account for regional differences in humidity, we utilized a partitional clustering algorithm with dynamic time warping (DTW) similarity measurements (*34*) to classify the absolute humidity profile for all observed counties into ten exclusive clusters. Clustering allowed us to designate groups of counties based on temporal, climatological regimes and to stratify different absolute humidity patterns, which reduces group-level effects and enhances the independence of the data points. The DTW clustering of absolute humidity was conducted on a larger set of 3,137 counties. In the regression analysis, we only examined counties that had more than twenty cumulative confirmed cases by September 29^th^, 2020, or a population of more than 50,000 people. We excluded any days that had fewer than 20 cumulative confirmed cases within each county because early transmission dynamics had a high rate of undetected cases (*35*), making the data unreliable for this analysis. The final dataset used in the regression analysis included 987 counties. We assessed the results of the model over three time periods in 2020: (1) the entire duration of the dataset (March 10 to Sept 29), (2) spring when humidity increases (March 10 to June 30), and (3) the summer months when humidity is relatively steady (July 1 to Sept 29).

For each humidity cluster, we conducted three multivariate regressions using a generalized additive model (GAM) that assessed the time-weighted association between absolute humidity and non-essential visits with the number of new coronavirus cases (Eqns. 1-3). GAMs are semiparametric models that have been used extensively in assessing impacts of climatic variables and health outcomes (*36, 37*) and allowed us to estimate a functional variability with basis splines (*38*). The model we used is described in the following equation,

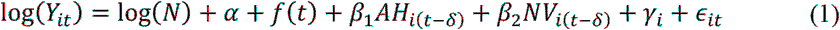

where *Y*_*it*_, is the number of daily COVID-19 cases for county *i* at time *t*, log(*N*) is an offset term to control for population-size, and *α* is the intercept. Absolute humidity, *AH*_*it-δ*_, and non-essential visitations, *NV*_*it-δ*_, were smoothed using a 7-day moving average and lagged by *δ* days. For our study, we assumed that *δ* was equal to 14 days, which is based on previous studies investigating lagged effects due to the incubation period of COVID-19 (*39*). Fixed effects, defined as *γ*_*i*_, are included for each county in our analysis to control for variations between counties. The operator *f*(·) is a spline basis function that is used to fit nonlinearities attributed to unobserved time-varying effects. Since we were predicting daily cases, we assumed the response variable is a Poisson distributed random variable with a log-transformed link function. Standard errors were calculated for the estimated linear coefficients *β*_1_ and *β*_*2*_.

To test for robustness, we conducted additional regressions on the absolute humidity and non-essential visits predictors individually with fixed effects for counties. Specifically, for each humidity cluster, we fitted a GAM with non-essential visitations and absolute humidity as linear predictors for new daily cases, as described in Equations (2) and (3).

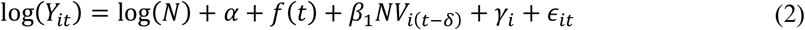

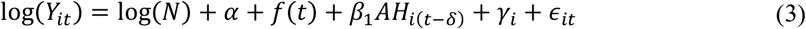

The multiple regression analyses were compared to demonstrate robustness in the coefficient estimates and the relative effects of absolute humidity and non-essential visitations. The analysis using GAM was conducted using the *mgcv* packages in R (Version 4.0.2).

## Results

Using the dynamic time warping (DTW) algorithm to characterize similarities of humidity profiles, we were able to partition all counties in our humidity dataset into ten exclusive groups. The clustering of humidity coincided with various geographic regions in the US (Figure 1A), which supports the notion that climate is spatially dependent. The cluster with the lowest average absolute humidity is Group 7 (398 counties), which is primarily located in the western region of the US. Group 6 (230 counties) had the highest average absolute humidity and is located on the southern coast bordering the Gulf of Mexico (see Figure 1B). Group 4 had the most varying geography, with counties from the West Coast, Midwest, and North East United States clustered together using the DTW algorithm.

**Figure 1.**
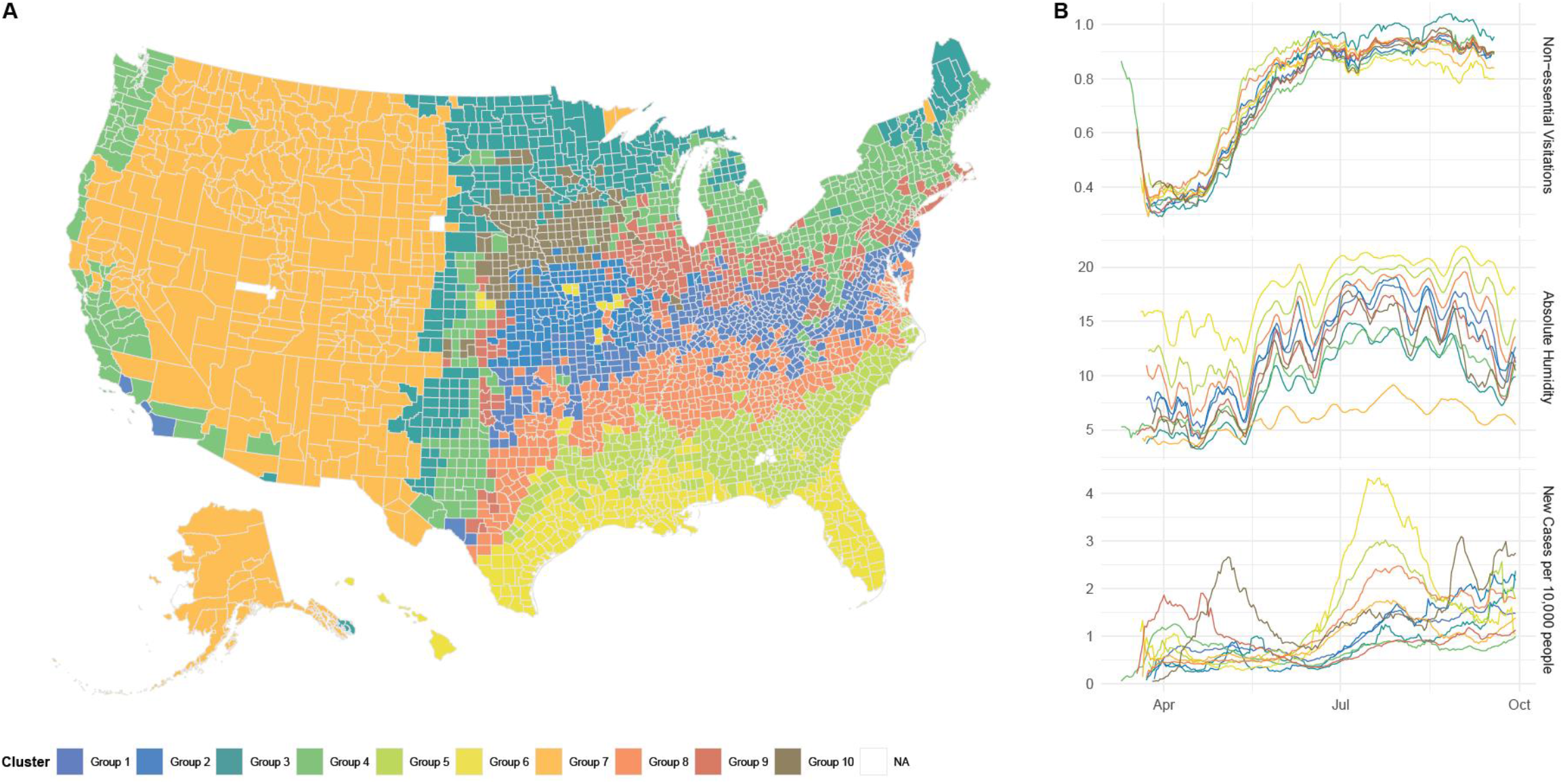
(A) Map of US Counties and their respective absolute humidity clusters. The clustering analysis was conducted using a partitional algorithm that utilized dynamic time warping (DTW) to measure similarity between absolute humidity profiles of 3,137 counties in the United States. Expectantly, the clustering of absolute humidity is related to the geography of the counties which serves as a proxy for regional weather patterns and different climatological regimes. (B) The cross-sectional smoothed mean of human encounter absolute humidity, and new case per 10,000 people trends for each cluster group of the 987 counties analyzed in the regression analysis.

The absolute humidity in counties that belonged to Groups 4 and 7 consistently had the largest negative effect on new cases for all three time periods (Tables 1-3). For the 7-month observation, Group 4 showed that a 1 g/m^3^ increase of absolute humidity resulted in a 0.21 decrease of new cases per day, while Group 7 had a 0.15 decrease in cases. When compared to the observations between March 10 and June 30, Group 4 still had a reduction of 0.22 new cases for each unit increase in absolute humidity, and Group 7 had a 0.18 decrease in cases. After July 1, absolute humidity had a relatively smaller impact on both groups, with Group 7 decreasing by 0.15 cases and Group 4 decreasing by 0.09 cases per day for each unit increase in absolute humidity. The cluster with the highest relative contribution to daily cases due to non-essential visitations for the entire six-month observation was Group 2 (Table 1). Group 9 had the largest increase in new cases due to an increase in non-essential visitations before July (Table 2), while Group 5 had the largest increase after July (Table 3). Generally, we see a stronger, positive association with non-essential visitations for all groups, except Group 9, for July to September (Table 3) than March to June (Table 2).

**Table 1.** GAM Regression against new cases from March 10, 2020 to September 29, 2020. The standard errors are shown in parenthesis.

|  | Group 1 | Group 2 | Group 3 | Group 4 | Group 5 | Group 6 | Group 7 | Group 8 | Group 9 | Group 10 |
| --- | --- | --- | --- | --- | --- | --- | --- | --- | --- | --- |
| *Predictors* | *Estimates* | *Estimates* | *Estimates* | *Estimates* | *Estimates* | *Estimates* | *Estimates* | *Estimates* | *Estimates* | *Estimates* |
| Intercept | -7.292 ***<br>(0.018) | -10.406 ***<br>(0.061) | -12.889 ***<br>(0.213) | -7.991 ***<br>(0.013) | -10.152 ***<br>(0.037) | -9.527 ***<br>(0.030) | -8.223 ***<br>(0.021) | -8.570 ***<br>(0.037) | -9.443 ***<br>(0.028) | -8.168 ***<br>(0.058) |
| Absolute Humidity<sup>†</sup><br>(14-day Lag) | **-0.114 \*\*\***<br>**(0.001)** | **-0.013 \*\*\***<br>**(0.002)** | **-0.028 \*\*\***<br>**(0.002)** | **-0.242 \*\*\***<br>**(0.001)** | 0.022 ***<br>(0.001) | **-0.006 \*\*\***<br>**(0.001)** | **-0.160 \*\*\***<br>**(0.001)** | **-0.011 \*\*\***<br>**(0.001)** | **-0.119 \*\*\***<br>**(0.001)** | **-0.006 \*\***<br>**(0.002)** |
| Non-essential Visitations<br>(14-day Lag) | -0.090 ***<br>(0.012) | 1.376 ***<br>(0.067) | 0.353 ***<br>(0.052) | 0.827 ***<br>(0.012) | 1.077 ***<br>(0.023) | 0.282 ***<br>(0.014) | -0.195 ***<br>(0.016) | -0.364 ***<br>(0.013) | 0.895 ***<br>(0.012) | -0.393 ***<br>(0.057) |
| spline(time) | 8.992 *** | 8.937 *** | 8.982 *** | 8.999 *** | 8.987 *** | 8.995 *** | 8.994 *** | 8.977 *** | 8.998 *** | 8.965 *** |
| Observations | 19834 | 4678 | 6020 | 38602 | 16301 | 16264 | 14670 | 22595 | 21067 | 3154 |
| R<sup>2</sup> | 0.569 | 0.638 | 0.625 | 0.655 | 0.365 | 0.629 | 0.772 | 0.556 | 0.675 | 0.457 |

\* $p < 0.05$ \*\* $p < 0.01$ \*\*\* $p < 0.001$
<sup>†</sup> Negative coefficient estimates are bolded
<sup>‡</sup> Estimated degree of freedom for spline basis function are listed

**Table 2.** GAM Regression against new cases from March 10, 2020 to June 30, 2020. The standard errors are shown in parenthesis.

|  | Group 1 | Group 2 | Group 3 | Group 4 | Group 5 | Group 6 | Group 7 | Group 8 | Group 9 | Group 10 |
| --- | --- | --- | --- | --- | --- | --- | --- | --- | --- | --- |
| *Predictors* | *Estimates* | *Estimates* | *Estimates* | *Estimates* | *Estimates* | *Estimates* | *Estimates* | *Estimates* | *Estimates* | *Estimates* |
| Intercept | -8.398 ***<br>(0.033) | -12.221 ***<br>(0.230) | -10.524 ***<br>(1.008) | -8.902 ***<br>(0.024) | -9.228 ***<br>(0.058) | -12.094 ***<br>(0.060) | -10.061 ***<br>(0.057) | -9.806 ***<br>(0.083) | -11.987 ***<br>(0.036) | -7.669 ***<br>(0.096) |
| Absolute Humidity<sup>†</sup><br>(14-day Lag) | **-0.123** ***<br>(0.002) | **-0.017** **<br>(0.006) | **-0.000**<br>(0.006) | **-0.254** ***<br>(0.001) | **-0.060** ***<br>(0.002) | 0.054 ***<br>(0.002) | **-0.194** ***<br>(0.002) | **-0.017** ***<br>(0.001) | 0.035 ***<br>(0.002) | 0.034 ***<br>(0.007) |
| Non-essential Visitations<br>(14-day Lag) | 0.854 ***<br>(0.035) | 1.422 ***<br>(0.128) | -0.614 ***<br>(0.100) | 0.993 ***<br>(0.024) | 0.976 ***<br>(0.036) | 0.585 ***<br>(0.029) | -0.223 ***<br>(0.035) | -0.183 ***<br>(0.029) | 3.110 ***<br>(0.027) | -1.849 ***<br>(0.115) |
| spline(time)<sup>‡</sup> | 8.989 *** | 8.924 *** | 8.821 *** | 8.996 *** | 8.974 *** | 8.981 *** | 8.971 *** | 8.942 *** | 8.996 *** | 8.645 *** |
| Observations | 8870 | 2103 | 2537 | 18573 | 7622 | 7852 | 6630 | 10532 | 9986 | 1445 |
| R<sup>2</sup> | 0.651 | 0.583 | 0.743 | 0.652 | 0.601 | 0.713 | 0.749 | 0.664 | 0.756 | 0.473 |

\* $p < 0.05$ \*\* $p < 0.01$ \*\*\* $p < 0.001$
<sup>†</sup> Negative coefficient estimates are bolded
<sup>‡</sup> Estimated degree of freedom for spline basis function are listed

**Table 3.** GAM Regression against new cases from July 1, 2020 to September 29, 2020. The standard errors are shown in parenthesis.

|  | Group 1 | Group 2 | Group 3 | Group 4 | Group 5 | Group 6 | Group 7 | Group 8 | Group 9 | Group 10 |
| --- | --- | --- | --- | --- | --- | --- | --- | --- | --- | --- |
| *Predictors* | *Estimates* | *Estimates* | *Estimates* | *Estimates* | *Estimates* | *Estimates* | *Estimates* | *Estimates* | *Estimates* | *Estimates* |
| Intercept | -10.444 ***<br>(0.046) | -12.546 ***<br>(0.094) | -14.296 ***<br>(0.266) | -9.361 ***<br>(0.028) | -12.846 ***<br>(0.058) | -8.502 ***<br>(0.050) | -7.736 ***<br>(0.034) | -10.581 ***<br>(0.065) | -11.816 ***<br>(0.073) | -12.645 ***<br>(0.126) |
| Absolute Humidity<sup>†</sup><br>(14-day Lag) | **-0.007** ***<br>(0.001) | 0.023 ***<br>(0.003) | **-0.023** ***<br>(0.003) | **-0.095** ***<br>(0.001) | 0.114 ***<br>(0.002) | **-0.051** ***<br>(0.001) | **-0.164** ***<br>(0.001) | 0.012 ***<br>(0.001) | **-0.026** ***<br>(0.002) | 0.016 ***<br>(0.003) |
| Non-essential Visitations<br>(14-day Lag) | 2.060 ***<br>(0.040) | 4.541 ***<br>(0.101) | 1.463 ***<br>(0.115) | 2.183 ***<br>(0.032) | 2.329 ***<br>(0.035) | 0.674 ***<br>(0.022) | 0.350 ***<br>(0.036) | 1.427 ***<br>(0.038) | 1.555 ***<br>(0.044) | 3.790 ***<br>(0.112) |
| spline(time)<sup>‡</sup> | 8.803 *** | 8.955 *** | 8.854 *** | 8.970 *** | 8.987 *** | 8.997 *** | 8.970 *** | 8.915 *** | 8.909 *** | 8.947 *** |
| Observations | 10964 | 2575 | 3483 | 20029 | 8679 | 8412 | 8040 | 12063 | 11081 | 1709 |
| R<sup>2</sup> | 0.650 | 0.647 | 0.607 | 0.727 | 0.352 | 0.619 | 0.792 | 0.541 | 0.816 | 0.556 |

\* $p < 0.05$ \*\* $p < 0.01$ \*\*\* $p < 0.001$
<sup>†</sup> Negative coefficient estimates are bolded
<sup>‡</sup> Estimated degree of freedom for spline basis function are listed

The coefficient estimates and fitted plots for the additional GAM to show robustness are shown in the supplementary appendix (Tables S1-S10). For all models, the estimated degrees of freedom for the time-varying basis spline were approximately 8 (see Figures S2 in supplementary appendix for fits). The coefficient estimates for Groups 4 and 7 were consistently the largest for those additional fits, indicating robustness in the results.

## Discussion

The approaching winter in the United States has been of great concern because of the link between wintertime and increasing transmission of respiratory viruses such as influenza and the other pandemic coronaviruses, MERS-CoV and SARS-CoV-1 (*9, 40*–*43*). SARS-CoV-2 is a novel human virus, and there remains great uncertainty as to the extent that disease transmission will increase over the next several months. Here we found that the relative effect of absolute humidity on transmissions has so far been significant and was greatest in the Western, Midwest, and Northeast regions of the United States, which were clustered into the driest climatological regimes. This study outcome supports the notion that absolute humidity affects transmission risk of respiratory viruses in regions that are arid and dry (*44*). However, this effect was less noticeable for more humid regions, such as the coastal counties and the southern regions of the US (Figure 2). We also found that non-essential visitations had a stronger, more positive impact on emergence of cases during the summer months. This suggests that an increase in contacts due to human mobility had a relatively more extensive role in case growth when absolute humidity is steady and higher.

**Figure 2.**
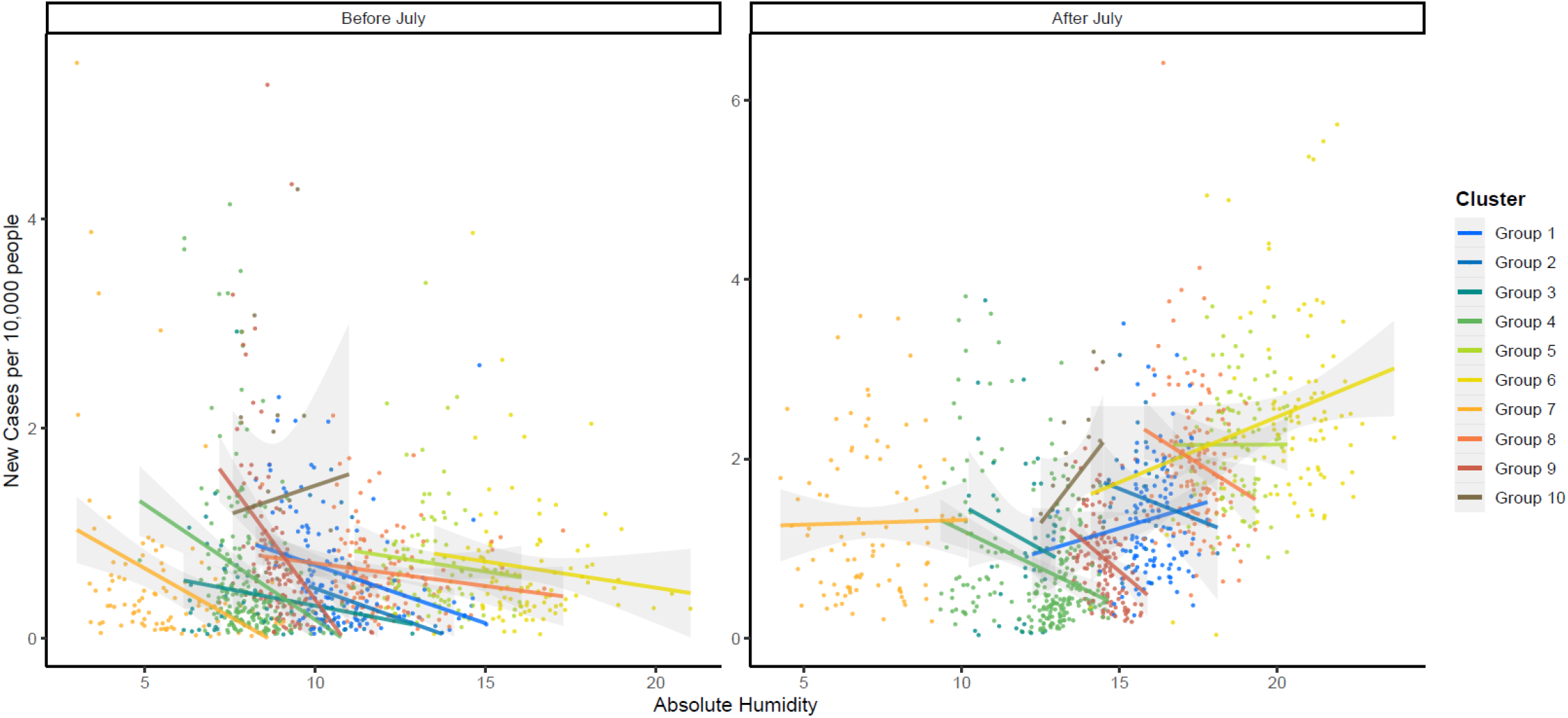
The average daily new cases plotted against average absolute humidity of 987 counties for March 10 to June 30 (*left*) and July 1 to September 29 (*right*). For each cluster group, we added a simple linear trend line with shaded standard errors. We see that within each humidity cluster before July, the absolute humidity in regions with lower humidity, i.e. Groups 6 and 7, has a larger negative slope correlation; however, after July when humidity patterns are stable, this relationship is less noticeable without considering the temporal relationships. The wetter regions also experience less impact from absolute humidity.

As the absolute humidity decreases during winter seasons, this suggests that COVID-19 may increase significantly over the next couple months. Increasing COVID-19 cases will pose an issue for many healthcare systems, which in normal years are typically stretched thin from regular seasonal infections, such as influenza. Furthermore, seasonal changes in human behavior may also impact the number of new cases and hospitalizations since people are more likely to occupy indoor spaces for longer durations when outdoor temperatures decrease, thus increasing the risk of transmission. With lower absolute humidity levels, relative humidity indoors will also be lower, causing higher susceptibility to airborne diseases (*45*).

The mechanisms through which changes in absolute humidity might affect transmission of respiratory viruses are not well understood. Several studies have shown that as absolute humidity falls, survival times for enveloped viruses increases, including other coronaviruses (*9, 20, 46*), though there are some suggestions that these effects may be nonlinear (*47*). Our findings supported this nonlinear relationship since the log-linear effects between humidity and case growth varied between climatological regimes. One potential factor may be that increasing humidity results in degradation of the capsid envelope leading to viral degradation (*46, 48*). Another possibility may be that higher absolute humidity reduces the virus’s binding capacity, reducing the potential infectivity (*48*). Further studies are needed to refine the mechanisms by which humidity changes may alter transmission of SARS-CoV-2 and the related functional form. However, the results of these prior studies and the association found in the current study suggest that increased humidification of indoor air in high transmission settings may help decrease the impact of COVID-19.

While our findings suggest that declines in absolute humidity are likely to contribute to increased transmission of SARS-CoV-2, our findings also demonstrate the importance of contact and movement of individuals. For example, in the 2009 influenza pandemic, the increased contact patterns that occur in the fall likely combined with falling humidity to drive transmission, which resulted in the peak of infections occurring significantly earlier than other years. Given the uncertainty of the nonlinear effects of humidity on transmission, continued restrictions on large gatherings and mandates for face-coverings are likely crucial in the coming months to reducing the toll of the pandemic. In addition, the uncertainty regarding the role of children in transmission (*49*–*51*) suggests that caution in opening schools is warranted as the potential for transmission increases, which contributes to seasonality effects. While studies linking schools to outbreaks to date have been limited, few have occurred during the winter when transmission may be higher. Studies assessing infections of children are urgently needed in areas where schools have opened.

Studies of prior viruses and preliminary studies of the SARS-CoV-2 virus underpin the theoretical connection between humidity and transmission; however, limitations remain in assessing this association. One limitation of this study includes the spatial coupling of populations due to travel and commute, which may alter the transmission patterns within counties. Spatial clustering of the analysis by trends in humidity should reduce this effect somewhat. A second limitation is an intra-county heterogeneity of the population (i.e., income, demographics, and social status), which we attempted to control for with fixed effects in our model. Third, as with most COVID-19 analyses on retrospective data, heterogeneity in testing at the county level likely results in differences in the correlation between reported results and actual cases. Fourth, other exogenous events, such as evacuation-related activities related to natural disasters or mass-gatherings in several cities this summer (*52*), may bias the connection between humidity and transmission. Finally, there may also be time-varying trends in other factors that affect case detection rates, though GAMs reduce the potential impact of these factors as they account for time-varying trends related to unobservable factors external to the independent variables in the model.

The upcoming winter in the United States and other temperate regions in the northern hemisphere are likely to increase transmission of SARS-CoV-2 due to falling humidity. In the US, these impacts are likely to be largest in the Western, Midwest, and Northeast regions of the United States that have lower humidity trends relative to the rest of the country. Given that much of our understanding of transmission has occurred during periods of higher humidity, significant uncertainty remains as to the impact of this increased transmission on morbidity and mortality of COVID-19. However, underestimating the potential for increased transmission could lead to needless deaths, particularly in nursing homes and long-term care facilities, where the possibility exists that infection control procedures in the summer may not be adequate in the winter and limits on visitors and regular screening of staff should be instituted sooner rather than later.

## Data Availability

The data that support the findings of this study are openly available through the Johns Hopkins Center for Systems Science and Engineering, Unacast Social Distancing Scorecard, and NOAA National Centers for Environmental Information.

## Acknowledgments

This work was funded by the Centers for Disease Control and Prevention (CDC) MInD-Healthcare Program (Grant Numbers U01CK000589, 1U01CK000536, and contract number 75D30120P07912). The funders had no role in the design, data collection and analysis, decision to publish, or preparation of the manuscript.

## Notes

### Competing Interest Statement

The authors have declared no competing interest.

### Author Declarations

Exempted from IRB oversight as the study is based on published secondary data.

