## Supplementary Appendix for "Investigating the effects of absolute humidity and human encounters on transmission of COVID-19 in the United States"

**Table S1.** GAM Regression against new cases for Group 1. The standard errors are shown in parenthesis.

| | **Both (March-Sept)** | **Absolute Humidity Model (March-Sept)** | **Non-essential Visitations (March-Sept)** | **Both (March-June)** | **Absolute Humidity Model (March-June)** | **Non-essential Visitations (March-June)** | **Both (July-Sept)** | **Absolute Humidity Model (July-Sept)** | **Non-essential Visitations (July-Sept)** |
|---|---|---|---|---|---|---|---|---|---|
| *Predictors* | *Log-Mean* | *Log-Mean* | *Log-Mean* | *Log-Mean* | *Log-Mean* | *Log-Mean* | *Log-Mean* | *Log-Mean* | *Log-Mean* |
| Intercept | -7.292 *** (0.018) | -7.351 *** (0.016) | -8.681 *** (0.015) | -8.398 *** (0.033) | -7.861 *** (0.025) | -9.620 *** (0.029) | -10.444 *** (0.046) | -8.597 *** (0.028) | -10.584 *** (0.038) |
| Absolute Humidity (14-day Lag) | -0.114 *** (0.001) | -0.115 *** (0.001) | | -0.123 *** (0.002) | -0.121 *** (0.002) | | -0.007 *** (0.001) | -0.011 *** (0.001) | |
| Non-essential Visitations (14-day Lag) | -0.090 *** (0.012) | | -0.331 *** (0.012) | 0.854 *** (0.035) | | 0.764 *** (0.034) | 2.060 *** (0.040) | | 2.083 *** (0.040) |
| spline(time) | 8.992 *** (NA) | 8.992 *** (NA) | 8.992 *** (NA) | 8.989 *** (NA) | 8.987 *** (NA) | 8.985 *** (NA) | 8.803 *** (NA) | 8.726 *** (NA) | 8.780 *** (NA) |
| Observations | 19834 | 19834 | 20196 | 8870 | 8870 | 9106 | 10964 | 10964 | 11090 |
| $R^2$ | 0.569 | 0.569 | 0.517 | 0.651 | 0.648 | 0.617 | 0.650 | 0.636 | 0.646 |

* *p<0.05* ** *p<0.01* *** *p<0.001*

**Table S2.** GAM Regression against new cases for Group 2. The standard errors are shown in parenthesis.

| *Predictors* | **Both (March-Sept)** *Estimates* | **Absolute Humidity Model (March-Sept)** *Estimates* | **Non-essential Visitations (March-Sept)** *Estimates* | **Both (March-June)** *Estimates* | **Absolute Humidity Model (March-June)** *Estimates* | **Non-essential Visitations (March-June)** *Estimates* | **Both (July-Sept)** *Estimates* | **Absolute Humidity Model (July-Sept)** *Estimates* | **Non-essential Visitations (July-Sept)** *Estimates* |
|---|---|---|---|---|---|---|---|---|---|
| Intercept | -10.406 *** (0.061) | -9.567 *** (0.045) | -10.639 *** (0.052) | -12.221 *** (0.230) | -11.372 *** (0.216) | -12.068 *** (0.166) | -12.546 *** (0.094) | -9.203 *** (0.056) | -12.123 *** (0.077) |
| Absolute Humidity (14-day Lag) | -0.013 *** (0.002) | -0.017 *** (0.002) | | -0.017 ** (0.006) | -0.023 *** (0.006) | | 0.023 *** (0.003) | 0.008 ** (0.003) | |
| Non-essential Visitations (14-day Lag) | 1.376 *** (0.067) | | 1.387 *** (0.066) | 1.422 *** (0.128) | | 1.398 *** (0.126) | 4.541 *** (0.101) | | 4.414 *** (0.100) |
| spline(time) | 8.937 *** | 8.931 *** | 8.940 *** | 8.924 *** | 8.885 *** | 8.935 *** | 8.955 *** | 8.861 *** | 8.951 *** |
| Observations | 4678 | 4678 | 4765 | 2103 | 2103 | 2152 | 2575 | 2575 | 2613 |
| $R^2$ | 0.638 | 0.634 | 0.639 | 0.583 | 0.569 | 0.585 | 0.647 | 0.629 | 0.648 |

* *p<0.05* ** *p<0.01* *** *p<0.001*

**Table S3.** GAM Regression against new cases for Group 3. The standard errors are shown in parenthesis.

| | **Both (March-Sept)** | **Absolute Humidity Model (March-Sept)** | **Non-essential Visitations (March-Sept)** | **Both (March-June)** | **Absolute Humidity Model (March-June)** | **Non-essential Visitations (March-June)** | **Both (July-Sept)** | **Absolute Humidity Model (July-Sept)** | **Non-essential Visitations (July-Sept)** |
|---|---|---|---|---|---|---|---|---|---|
| *Predictors* | *Estimates* | *Estimates* | *Estimates* | *Estimates* | *Estimates* | *Estimates* | *Estimates* | *Estimates* | *Estimates* |
| Intercept | -12.889 *** (0.213) | -12.406 *** (0.201) | -13.193 *** (0.212) | -10.524 *** (1.008) | -11.276 *** (1.001) | -10.524 *** (1.007) | -14.296 *** (0.266) | -12.142 *** (0.206) | -14.619 *** (0.263) |
| Absolute Humidity (14-day Lag) | -0.028 *** (0.002) | -0.029 *** (0.002) | | -0.000 (0.006) | 0.004 (0.005) | | -0.023 *** (0.003) | -0.025 *** (0.003) | |
| Non-essential Visitations (14-day Lag) | 0.353 *** (0.052) | | 0.394 *** (0.051) | -0.614 *** (0.100) | | -0.614 *** (0.099) | 1.463 *** (0.115) | | 1.510 *** (0.114) |
| spline(time) | 8.982 *** | 8.984 *** | 8.985 *** | 8.821 *** | 8.837 *** | 8.845 *** | 8.854 *** | 8.775 *** | 8.853 *** |
| Observations | 6020 | 6020 | 6035 | 2537 | 2537 | 2544 | 3483 | 3483 | 3491 |
| $R^2$ | 0.625 | 0.625 | 0.624 | 0.743 | 0.744 | 0.743 | 0.607 | 0.605 | 0.606 |

* *p<0.05* ** *p<0.01* *** *p<0.001*

**Table S4.** GAM Regression against new cases for Group 4. The standard errors are shown in parenthesis.

| | **Both (March-Sept)** | **Absolute Humidity Model (March-Sept)** | **Non-essential Visitations (March-Sept)** | **Both (March-June)** | **Absolute Humidity Model (March-June)** | **Non-essential Visitations (March-June)** | **Both (July-Sept)** | **Absolute Humidity Model (July-Sept)** | **Non-essential Visitations (July-Sept)** |
|---|---|---|---|---|---|---|---|---|---|
| *Predictors* | *Estimates* | *Estimates* | *Estimates* | *Estimates* | *Estimates* | *Estimates* | *Estimates* | *Estimates* | *Estimates* |
| Intercept | -7.991 *** (0.013) | -7.567 *** (0.012) | -8.261 *** (0.012) | -8.902 *** (0.024) | -8.347 *** (0.019) | -9.608 *** (0.022) | -9.361 *** (0.028) | -7.890 *** (0.017) | -10.391 *** (0.024) |
| Absolute Humidity (14-day Lag) | -0.242 *** (0.001) | -0.228 *** (0.000) | | -0.254 *** (0.001) | -0.241 *** (0.001) | | -0.095 *** (0.001) | -0.106 *** (0.001) | |
| Non-essential Visitations (14-day Lag) | 0.827 *** (0.012) | | -1.243 *** (0.010) | 0.993 *** (0.024) | | 0.215 *** (0.024) | 2.183 *** (0.032) | | 2.706 *** (0.032) |
| spline(time) | 8.999 *** | 8.998 *** | 8.998 *** | 8.996 *** | 8.996 *** | 8.991 *** | 8.970 *** | 8.980 *** | 8.959 *** |
| Observations | 38602 | 38611 | 39037 | 18573 | 18582 | 18674 | 20029 | 20029 | 20363 |
| $R^2$ | 0.655 | 0.655 | 0.578 | 0.652 | 0.655 | 0.566 | 0.727 | 0.720 | 0.724 |

* *p<0.05* ** *p<0.01* *** *p<0.001*

**Table S5.** GAM Regression against new cases for Group 5. The standard errors are shown in parenthesis.

| | **Both (March-Sept)** | **Absolute Humidity Model (March-Sept)** | **Non-essential Visitations (March-Sept)** | **Both (March-June)** | **Absolute Humidity Model (March-June)** | **Non-essential Visitations (March-June)** | **Both (July-Sept)** | **Absolute Humidity Model (July-Sept)** | **Non-essential Visitations (July-Sept)** |
|---|---|---|---|---|---|---|---|---|---|
| *Predictors* | *Log-Mean* | *Log-Mean* | *Log-Mean* | *Log-Mean* | *Log-Mean* | *Log-Mean* | *Log-Mean* | *Log-Mean* | *Log-Mean* |
| Intercept | -10.152 *** (0.037) | -9.068 *** (0.028) | -9.765 *** (0.032) | -9.228 *** (0.058) | -8.458 *** (0.050) | -9.943 *** (0.053) | -12.846 *** (0.058) | -10.114 *** (0.041) | -10.403 *** (0.046) |
| Absolute Humidity (14-day Lag) | 0.022 *** (0.001) | 0.018 *** (0.001) | | -0.060 *** (0.002) | -0.057 *** (0.002) | | 0.114 *** (0.002) | 0.099 *** (0.002) | |
| Non-essential Visitations (14-day Lag) | 1.077 *** (0.023) | | 1.027 *** (0.023) | 0.976 *** (0.036) | | 0.927 *** (0.036) | 2.329 *** (0.035) | | 2.001 *** (0.034) |
| spline(time) | 8.987 *** (NA) | 8.982 *** (NA) | 8.988 *** (NA) | 8.974 *** (NA) | 8.962 *** (NA) | 8.966 *** (NA) | 8.987 *** (NA) | 8.989 *** (NA) | 8.991 *** (NA) |
| Observations | 16301 | 16301 | 16760 | 7622 | 7622 | 7854 | 8679 | 8679 | 8906 |
| $R^2$ | 0.365 | 0.365 | 0.363 | 0.601 | 0.599 | 0.575 | 0.352 | 0.351 | 0.342 |

* *p<0.05* ** *p<0.01* *** *p<0.001*

**Table S6.** GAM Regression against new cases for Group 6. The standard errors are shown in parenthesis.

| | **Both (March-Sept)** | **Absolute Humidity Model (March-Sept)** | **Non-essential Visitations (March-Sept)** | **Both (March-June)** | **Absolute Humidity Model (March-June)** | **Non-essential Visitations (March-June)** | **Both (July-Sept)** | **Absolute Humidity Model (July-Sept)** | **Non-essential Visitations (July-Sept)** |
|---|---|---|---|---|---|---|---|---|---|
| *Predictors* | *Estimates* | *Estimates* | *Estimates* | *Estimates* | *Estimates* | *Estimates* | *Estimates* | *Estimates* | *Estimates* |
| Intercept | -9.527 *** (0.030) | -9.069 *** (0.020) | -9.664 *** (0.025) | -12.094 *** (0.060) | -11.364 *** (0.047) | -11.171 *** (0.053) | -8.502 *** (0.050) | -7.316 *** (0.031) | -9.525 *** (0.039) |
| Absolute Humidity (14-day Lag) | -0.006 *** (0.001) | -0.008 *** (0.001) | | 0.054 *** (0.002) | 0.052 *** (0.002) | | -0.051 *** (0.001) | -0.053 *** (0.001) | |
| Non-essential Visitations (14-day Lag) | 0.282 *** (0.014) | | 0.296 *** (0.014) | 0.585 *** (0.029) | | 0.527 *** (0.028) | 0.674 *** (0.022) | | 0.654 *** (0.022) |
| spline(time) | 8.995 *** | 8.995 *** | 8.996 *** | 8.981 *** | 8.980 *** | 8.985 *** | 8.997 *** | 8.997 *** | 8.996 *** |
| Observations | 16264 | 16264 | 16634 | 7852 | 7852 | 8046 | 8412 | 8412 | 8588 |
| $R^2$ | 0.629 | 0.628 | 0.628 | 0.713 | 0.711 | 0.706 | 0.619 | 0.617 | 0.617 |

* *p<0.05* ** *p<0.01* *** *p<0.001*

**Table S7.** GAM Regression against new cases for Group 7. The standard errors are shown in parenthesis.

| | **Both (March-Sept)** | **Absolute Humidity Model (March-Sept)** | **Non-essential Visitations (March-Sept)** | **Both (March-June)** | **Absolute Humidity Model (March-June)** | **Non-essential Visitations (March-June)** | **Both (July-Sept)** | **Absolute Humidity Model (July-Sept)** | **Non-essential Visitations (July-Sept)** |
|---|---|---|---|---|---|---|---|---|---|
| *Predictors* | *Estimates* | *Estimates* | *Estimates* | *Estimates* | *Estimates* | *Estimates* | *Estimates* | *Estimates* | *Estimates* |
| Intercept | -8.223 *** (0.021) | -8.367 *** (0.017) | -9.519 *** (0.020) | -10.061 *** (0.057) | -10.197 *** (0.053) | -10.837 *** (0.058) | -7.736 *** (0.034) | -7.466 *** (0.019) | -9.185 *** (0.033) |
| Absolute Humidity (14-day Lag) | -0.160 *** (0.001) | -0.159 *** (0.001) | | -0.194 *** (0.002) | -0.196 *** (0.002) | | -0.164 *** (0.001) | -0.163 *** (0.001) | |
| Non-essential Visitations (14-day Lag) | -0.195 *** (0.016) | | -0.030 (0.017) | -0.223 *** (0.035) | | -0.608 *** (0.039) | 0.350 *** (0.036) | | 0.388 *** (0.037) |
| spline(time) | 8.994 *** | 8.994 *** | 8.991 *** | 8.971 *** | 8.973 *** | 8.875 *** | 8.970 *** | 8.971 *** | 8.966 *** |
| Observations | 14670 | 14833 | 14729 | 6630 | 6710 | 6630 | 8040 | 8123 | 8099 |
| $R^2$ | 0.772 | 0.771 | 0.712 | 0.749 | 0.745 | 0.723 | 0.792 | 0.792 | 0.724 |

* *p<0.05*   ** *p<0.01*   *** *p<0.001*

**Table S8.** GAM Regression against new cases for Group 8. The standard errors are shown in parenthesis.

| | **Both (March-Sept)** | **Absolute Humidity Model (March-Sept)** | **Non-essential Visitations (March-Sept)** | **Both (March-June)** | **Absolute Humidity Model (March-June)** | **Non-essential Visitations (March-June)** | **Both (July-Sept)** | **Absolute Humidity Model (July-Sept)** | **Non-essential Visitations (July-Sept)** |
|---|---|---|---|---|---|---|---|---|---|
| *Predictors* | *Log-Mean* | *Log-Mean* | *Log-Mean* | *Log-Mean* | *Log-Mean* | *Log-Mean* | *Log-Mean* | *Log-Mean* | *Log-Mean* |
| Intercept | -8.570 *** (0.037) | -9.006 *** (0.033) | -8.265 *** (0.029) | -9.806 *** (0.083) | -10.000 *** (0.077) | -10.028 *** (0.081) | -10.581 *** (0.065) | -8.649 *** (0.040) | -9.982 *** (0.055) |
| Absolute Humidity (14-day Lag) | -0.011 *** (0.001) | -0.011 *** (0.001) | | -0.017 *** (0.001) | -0.016 *** (0.001) | | 0.012 *** (0.001) | 0.007 *** (0.001) | |
| Non-essential Visitations (14-day Lag) | -0.364 *** (0.013) | | -0.495 *** (0.013) | -0.183 *** (0.029) | | -0.134 *** (0.029) | 1.427 *** (0.038) | | 1.354 *** (0.037) |
| spline(time) | 8.977 *** (NA) | 8.979 *** (NA) | 8.978 *** (NA) | 8.942 *** (NA) | 8.947 *** (NA) | 8.905 *** (NA) | 8.915 *** (NA) | 8.903 *** (NA) | 8.941 *** (NA) |
| Observations | 22595 | 22595 | 23138 | 10532 | 10532 | 10756 | 12063 | 12063 | 12382 |
| $R^2$ | 0.556 | 0.556 | 0.556 | 0.664 | 0.664 | 0.657 | 0.541 | 0.540 | 0.540 |

* *p<0.05* ** *p<0.01* *** *p<0.001*

**Table S9.** GAM Regression against new cases for Group 9. The standard errors are shown in parenthesis.

| | **Both (March-Sept)** | **Absolute Humidity Model (March-Sept)** | **Non-essential Visitations (March-Sept)** | **Both (March-June)** | **Absolute Humidity Model (March-June)** | **Non-essential Visitations (March-June)** | **Both (July-Sept)** | **Absolute Humidity Model (July-Sept)** | **Non-essential Visitations (July-Sept)** |
|---|---|---|---|---|---|---|---|---|---|
| *Predictors* | *Estimates* | *Estimates* | *Estimates* | *Estimates* | *Estimates* | *Estimates* | *Estimates* | *Estimates* | *Estimates* |
| Intercept | -9.443 *** (0.028) | -8.711 *** (0.026) | -10.721 *** (0.026) | -11.987 *** (0.036) | -9.761 *** (0.030) | -11.712 *** (0.033) | -11.816 *** (0.073) | -10.149 *** (0.056) | -12.281 *** (0.068) |
| Absolute Humidity (14-day Lag) | -0.119 *** (0.001) | -0.117 *** (0.001) | | 0.035 *** (0.002) | 0.017 *** (0.002) | | -0.026 *** (0.002) | -0.032 *** (0.002) | |
| Non-essential Visitations (14-day Lag) | 0.895 *** (0.012) | | 0.840 *** (0.012) | 3.110 *** (0.027) | | 3.058 *** (0.027) | 1.555 *** (0.044) | | 1.619 *** (0.044) |
| spline(time) | 8.998 *** | 8.997 *** | 8.998 *** | 8.996 *** | 8.983 *** | 8.996 *** | 8.909 *** | 8.915 *** | 8.909 *** |
| Observations | 21067 | 21067 | 21418 | 9986 | 9986 | 10157 | 11081 | 11081 | 11261 |
| $R^2$ | 0.675 | 0.675 | 0.661 | 0.756 | 0.751 | 0.756 | 0.816 | 0.814 | 0.813 |

* *p<0.05* ** *p<0.01* *** *p<0.001*

**Table S10.** GAM Regression against new cases for Group 10. The standard errors are shown in parenthesis.

| | **Both (March-Sept)** | **Absolute Humidity Model (March-Sept)** | **Non-essential Visitations (March-Sept)** | **Both (March-June)** | **Absolute Humidity Model (March-June)** | **Non-essential Visitations (March-June)** | **Both (July-Sept)** | **Absolute Humidity Model (July-Sept)** | **Non-essential Visitations (July-Sept)** |
|---|---|---|---|---|---|---|---|---|---|
| *Predictors* | *Estimates* | *Estimates* | *Estimates* | *Estimates* | *Estimates* | *Estimates* | *Estimates* | *Estimates* | *Estimates* |
| Intercept | -8.168 *** (0.058) | -8.510 *** (0.030) | -8.231 *** (0.052) | -7.669 *** (0.096) | -8.972 *** (0.056) | -7.370 *** (0.078) | -12.645 *** (0.126) | -8.752 *** (0.051) | -12.460 *** (0.118) |
| Absolute Humidity (14-day Lag) | -0.006 ** (0.002) | -0.006 ** (0.002) | | 0.034 *** (0.007) | 0.044 *** (0.007) | | 0.016 *** (0.003) | 0.017 *** (0.003) | |
| Non-essential Visitations (14-day Lag) | -0.393 *** (0.057) | | -0.401 *** (0.057) | -1.849 *** (0.115) | | -1.901 *** (0.114) | 3.790 *** (0.112) | | 3.822 *** (0.112) |
| spline(time) | 8.965 *** | 8.969 *** | 8.968 *** | 8.645 *** | 8.854 *** | 8.461 *** | 8.947 *** | 8.951 *** | 8.947 *** |
| Observations | 3154 | 3154 | 3170 | 1445 | 1445 | 1445 | 1709 | 1709 | 1725 |
| $R^2$ | 0.457 | 0.458 | 0.457 | 0.473 | 0.464 | 0.472 | 0.556 | 0.531 | 0.555 |

* *p<0.05* ** *p<0.01* *** *p<0.001*

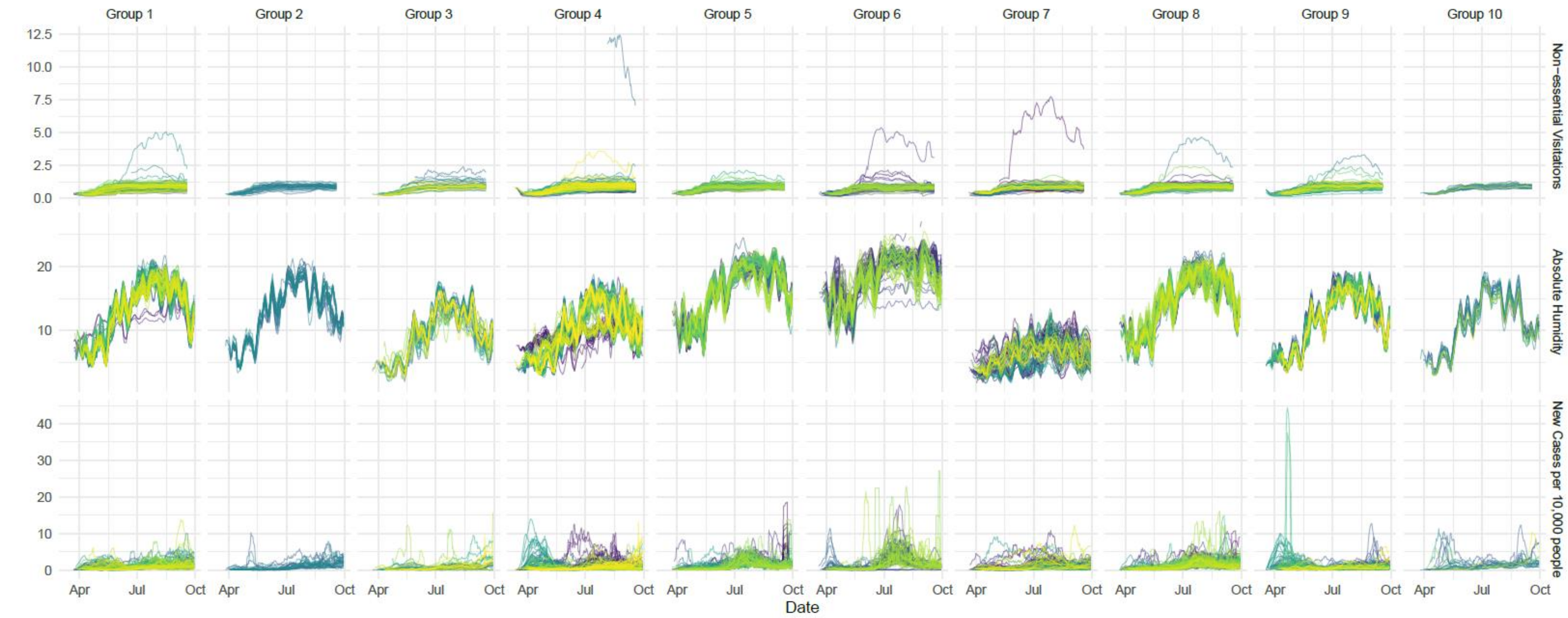


**Figure S1. Non-essential travel, absolute humidity, and daily case trends based on absolute humidity clusters.** All three series are based on a smoothed 7-day moving average.

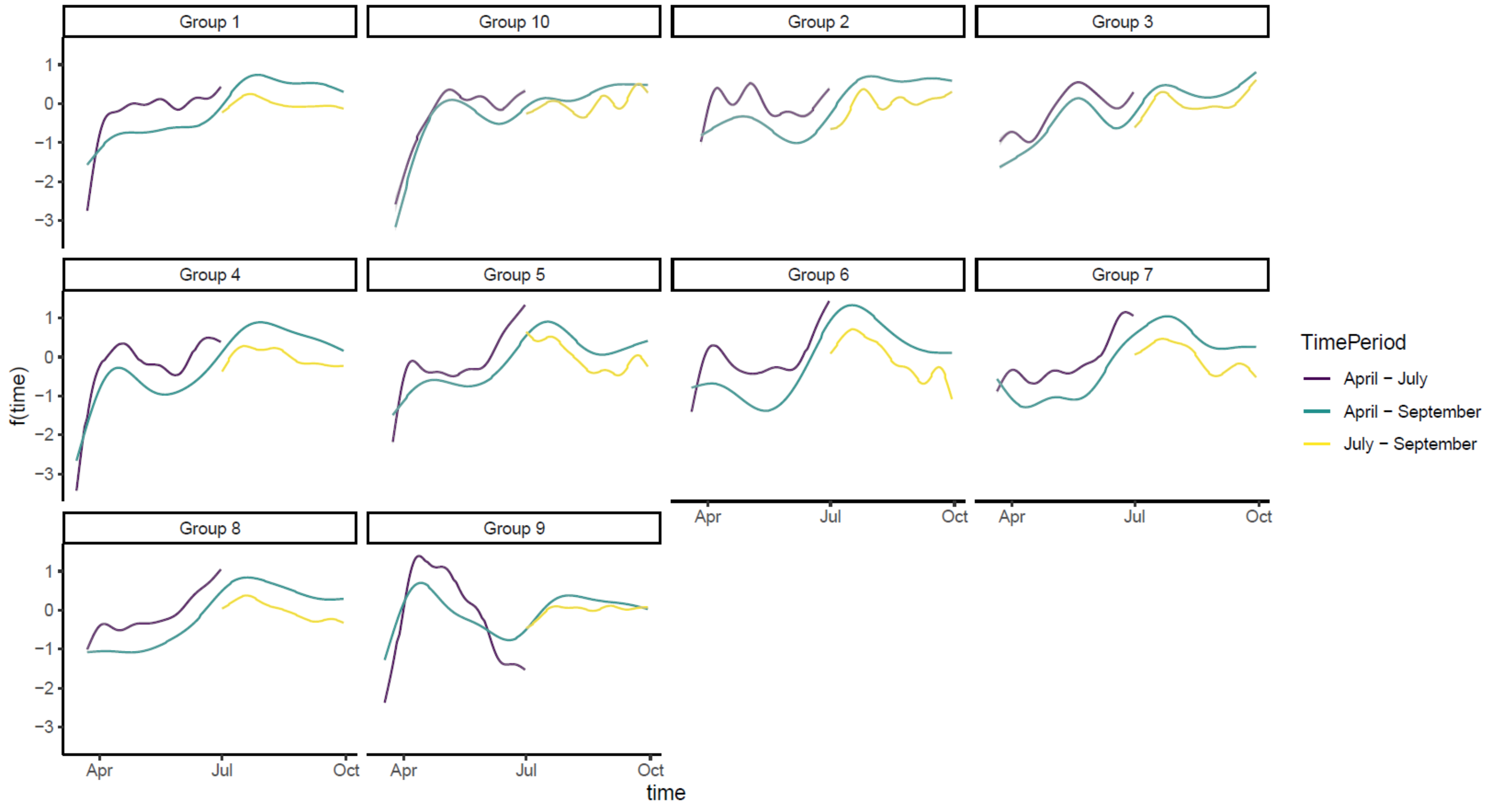


**Figure S2.** Spline fits from the GAM analysis described in Equation (1) for all three time periods. The fitted standard errors resulting from the residuals are shaded in grey.

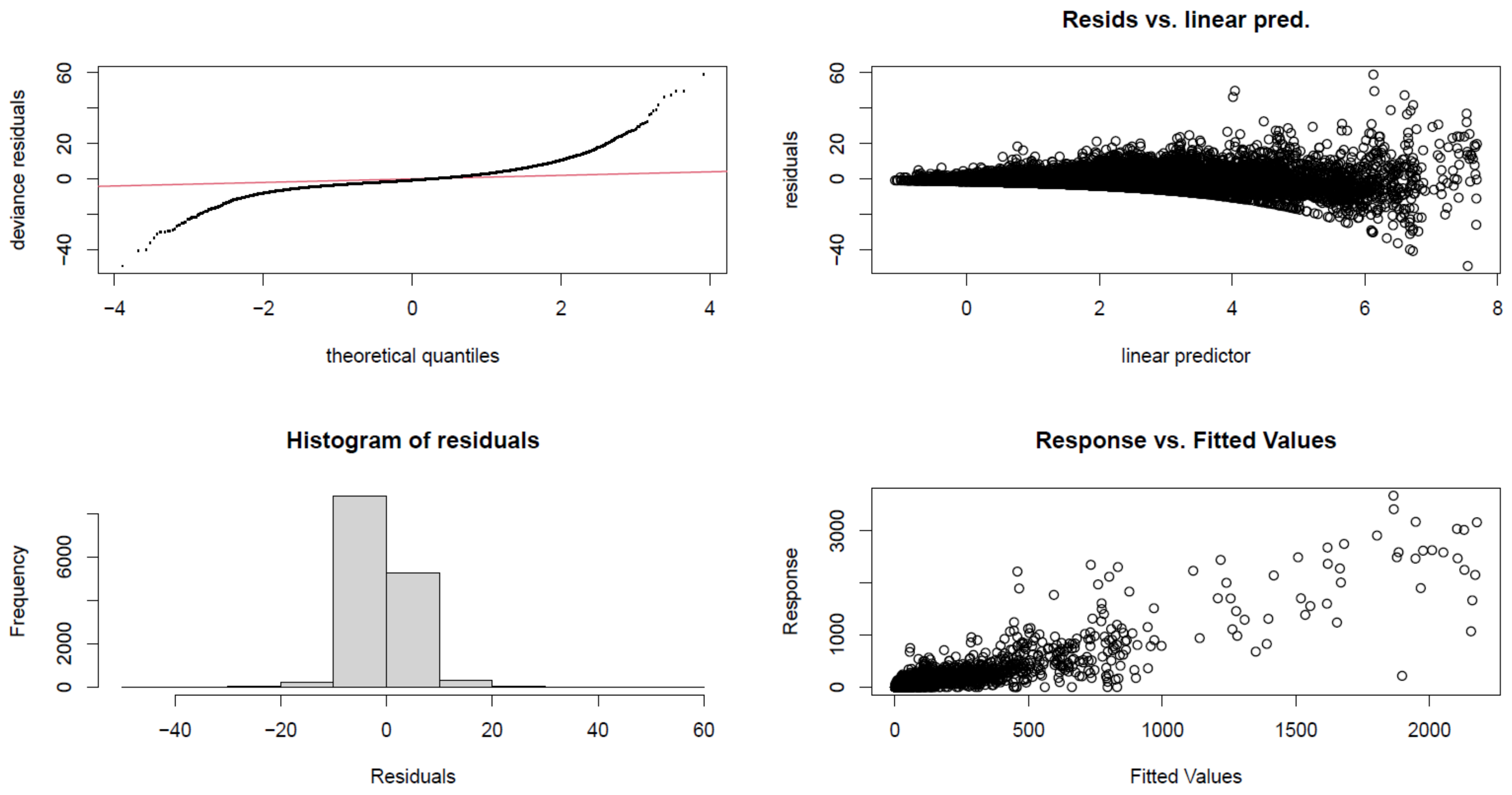


**Figure S3.** Residuals analysis for Group 7 cluster.

| Cluster | Non-essential visits (14-day lag) | Absolute Humidity (14-day lag) |
|---|---|---|
| Group 1 | 0.023 | 0.084 |
| Group 2 | -0.078 | -0.116 |
| Group 3 | -0.199 | -0.217 |
| Group 4 | -0.113 | -0.259 |
| Group 5 | -0.111 | 0.045 |
| Group 6 | 0.037 | 0.105 |
| Group 7 | -0.062 | -0.089 |
| Group 8 | -0.065 | 0.019 |
| Group 9 | -0.048 | -0.068 |
| Group 10 | 0.074 | -0.161 |

**Figure S4.** Pearson Correlation between daily new cases and predictor variables.
